# Femtomolar SARS-CoV-2 Antigen Detection Using the Microbubbling Digital Assay with Smartphone Readout Enables Antigen Burden Quantitation and Dynamics Tracking

**DOI:** 10.1101/2021.03.17.21253847

**Authors:** Hui Chen, Zhao Li, Sheng Feng, Anni Wang, Melissa Richard-Greenblatt, Emily Hutson, Stefen Andrianus, Laurel J. Glaser, Kyle G. Rodino, Jianing Qian, Dinesh Jayaraman, Ronald G. Collman, Abigail Glascock, Frederic D. Bushman, Jae Seung Lee, Sara Cherry, Alejandra Fausto, Susan R. Weiss, Hyun Koo, Patricia M. Corby, Una O’Doherty, Alfred L. Garfall, Dan T. Vogl, Edward A. Stadtmauer, Ping Wang

**Author notes:** Address correspondence to: Ping Wang, Department of Pathology and Laboratory Medicine, University of Pennsylvania, 3400 Spruce St., Founders 7.103, Philadelphia, PA. These two authors contributed equally (Hui Chen, Zhao Li).

## Abstract

**Background:** Little is known about the dynamics of SARS-CoV-2 antigen burden in respiratory samples in different patient populations at different stages of infection. Current rapid antigen tests cannot quantitate and track antigen dynamics with high sensitivity and specificity in respiratory samples.

**Methods:** We developed and validated an ultra-sensitive SARS-CoV-2 antigen assay with smartphone readout using the Microbubbling Digital Assay previously developed by our group, which is a platform that enables highly sensitive detection and quantitation of protein biomarkers. A computer vision-based algorithm was developed for microbubble smartphone image recognition and quantitation. A machine learning-based classifier was developed to classify the smartphone images based on detected microbubbles. Using this assay, we tracked antigen dynamics in serial swab samples from COVID patients hospitalized in ICU and immunocompromised COVID patients.

**Results:** The limit of detection (LOD) of the Microbubbling SARS-CoV-2 Antigen Assay was 0.5 pg/mL (10.6 fM) recombinant nucleocapsid (N) antigen or 4000 copies/mL inactivated SARS-CoV-2 virus in nasopharyngeal (NP) swabs, comparable to many rRT-PCR methods. The assay had high analytical specificity towards SARS-CoV-2. Compared to EUA-approved rRT-PCR methods, the Microbubbling Antigen Assay demonstrated a positive percent agreement (PPA) of 97% (95% confidence interval (CI), 92-99%) in symptomatic individuals within 7 days of symptom onset and positive SARS-CoV-2 nucleic acid results, and a negative percent agreement (NPA) of 97% (95% CI, 94-100%) in symptomatic and asymptomatic individuals with negative nucleic acid results. Antigen positivity rate in NP swabs gradually decreased as days-after-symptom-onset increased, despite persistent nucleic acid positivity of the same samples. The computer vision and machine learning-based automatic microbubble image classifier could accurately identify positives and negatives, based on microbubble counts and sizes. Total microbubble volume, a potential marker of antigen burden, correlated inversely with Ct values and days-after-symptom-onset. Antigen was detected for longer periods of time in immunocompromised patients with hematologic malignancies, compared to immunocompetent individuals. Simultaneous detectable antigens and nucleic acids may indicate the presence of replicating viruses in patients with persistent infections.

**Conclusions:** The Microbubbling SARS-CoV-2 Antigen Assay enables sensitive and specific detection of acute infections, and quantitation and tracking of antigen dynamics in different patient populations at various stages of infection. With smartphone compatibility and automated image processing, the assay is well-positioned to be adapted for point-of-care diagnosis and to explore the clinical implications of antigen dynamics in future studies.

## Introduction

A significant challenge in the SARS-CoV-2 pandemic is to develop sensitive, specific and easily accessible diagnostics to identify infections early, and to monitor infection progress in order to implement appropriate isolation and infection control procedures. Current diagnostic gold standard is real-time reverse transcription−polymerase chain reaction (rRT-PCR), with various methods demonstrating limit of detection (LOD) from 10^2^ to 10^5^ copies/mL according to manufacturer package inserts and reference panels ^1-3^. While many nucleic acid-based methods are sensitive enough to detect acute infections, persistent nucleic acid positivity after symptom resolution and disease recovery makes it challenging to determine the right level of infection control measures during patient care ^4^. Several lateral flow antigen assays have received FDA EUA approval. Due to the limit in analytical sensitivity, these tests generate more false negatives compared to nucleic acid tests, leading to concerns of missing acute infections ^5^. Furthermore, little is known regarding how antigen burden changes during the various stages of SARS-CoV-2 infection, and how this correlates with nucleic acid burden and infectivity in different patient populations. Therefore, highly sensitive, specific and convenient diagnostic methods that can detect acute infections, quantitate and track antigen dynamics during the disease course are highly desirable. In this study, we developed and validated a highly sensitive and specific SARS-CoV-2 antigen assay using smartphone readout, with computer vision image recognition and machine learning (ML) classification, for acute SARS-CoV-2 infection detection, antigen burden quantitation and tracking.

Nucleocapsid (N) protein is one of the most abundantly expressed proteins during the replication of SARS-CoV-2, and is evolutionally conservative with a low chance of mutations, and therefore widely used as the detection target in SARS-CoV-2 antigen assays.^6^ In order to detect acute infections at or before symptom onset, the key challenge is to have an assay sensitive enough to detect the very low concentrations of N in the early phase of infection. In this study, we use the Microbubbling Digital Assay previously developed by our group ^7^ to achieve high sensitivity SARS-CoV-2 antigen detection. In this technology, individual sandwich complexes formed between magnetic bead/target/PtNP are distributed in microwells in a microchip. Images of oxygen microbubbles generated through PtNP catalysis of H_2_O_2_ breakdown are captured using a smartphone camera as the detection method ^7^. We have previously shown that the Microbubbling Digital Assay can detect and quantitate prostate specific antigen (PSA) and beta human Chorionic Gonadotropin (βhCG) with high analytical sensitivity (limit of detection (LOD) of 2.1 fM for PSA and 0.034 mIU/mL for βhCG, several hundred folder higher sensitivity than current clinical methods) ^7^. We hereby take advantage of the features of the Microbubbling Digital Assay with directly visible signals, femtomolar analytical sensitivity, digital quantitation capability and compatibility with smartphone and artificial intelligence^7^, apply it to the detection of SARS-CoV-2 N antigen, and assess its performance in clinical swab samples. Using this assay, we tracked antigen dynamics in an ICU COVID inpatient population and an immunocompromised COVID patient population.

## Results

### Design and Characterization of the Microbubbling SARS-CoV-2 Antigen Assay

To repurpose the technology for the detection of the very low concentrations of SARS-CoV-2 N antigen during acute infections, we have designed the Microbubbling SARS-CoV-2 Antigen Assay as show in Figure 1. Nasopharyngeal (NP) swab, the same sample used in rRT-PCR methods, is eluted into saline or viral transport media, and treated with lysis buffer to release the N protein from the viruses. The released N protein is then detected using the Microbubbling assay, in which a monoclonal capture antibody is conjugated to the surface of magnetic microbeads, and a polyclonal detection antibody is biotinylated to bind to avidin coated platinum nanoparticles (PtNPs). When N protein is present, immunosandwich complexes form between the magnetic microbeads and PtNPs. After washing, the immunocomplexes are pulled down into a microwell array in the Microbubbling Chip via an external magnetic field, where microbubbles are generated through degradation of H_2_O_2_ catalyzed by PtNPs. The microbubble images, captured using a smartphone camera and a mobile microscope, are analyzed by computer vision and ML algorithms, which generate quantitative outputs (bubble size, number, total bubble volume) and classify the images as either SARS-CoV-2 positive or negative.

**Figure 1.**
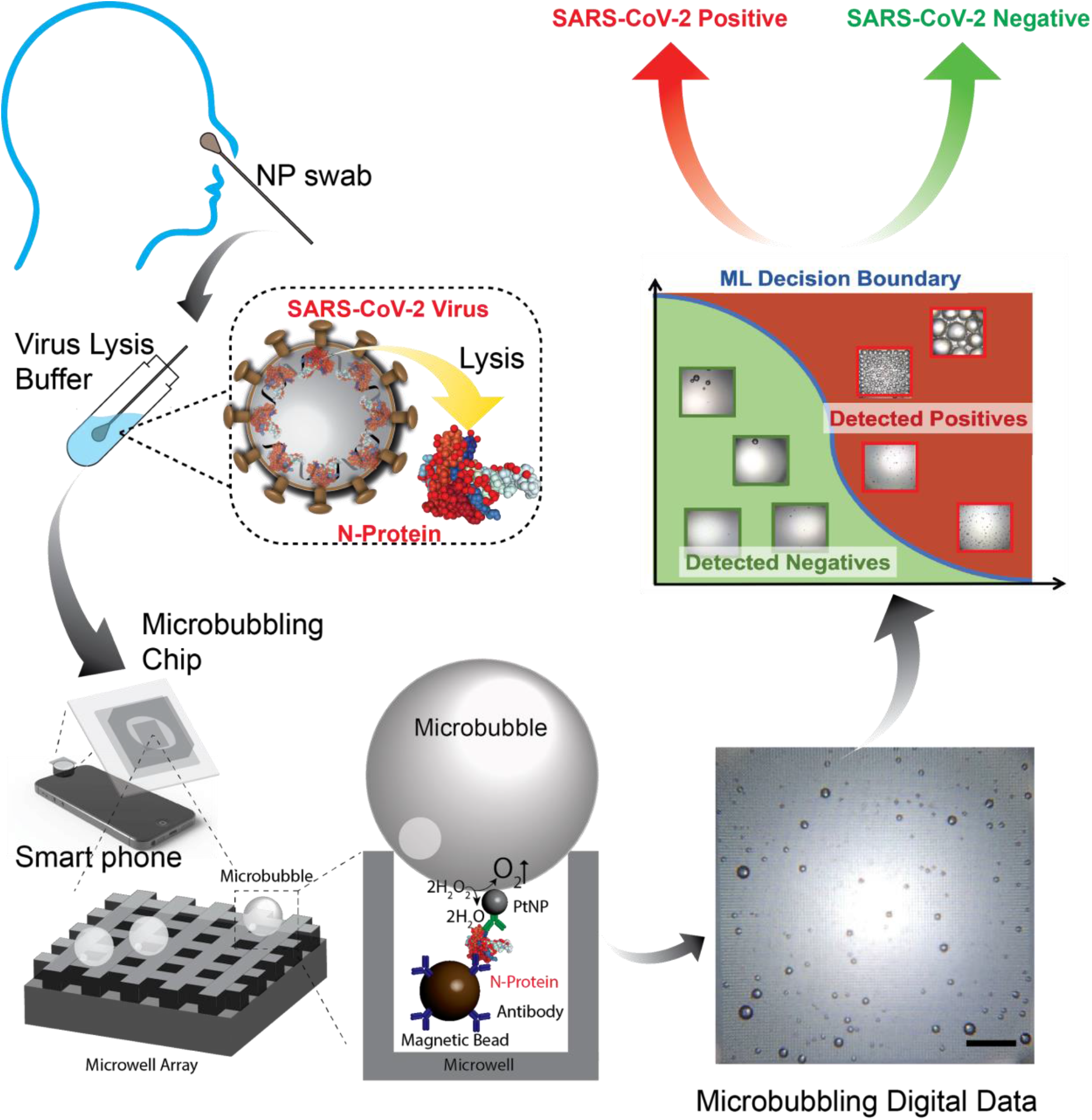
Schematic diagram of the procedures of the Microbubbling SARS-CoV-2 Antigen Assay. Nasopharyngeal (NP) swab eluant is first treated with lysis buffer to release N proteins from SARS-CoV-2 viruses. N protein is then detected by the smartphone-based microbubbling digital assay. The microbubble images are quantitated and classified as positive or negative by computer vision and ML algorithms.

To identify the best antibody pair to use in the Microbubbling SARS-CoV-2 assay, we screened different antibody pairs that bind the SARS-CoV-2 N protein with various capture/detection combinations (Supplementary Figure 1). The pair (N1 as capture antibody, Np as detection antibody) that generated the highest analytical sensitivity was chosen to design the assay (Figure 1). To assess the intrinsic analytical sensitivity of the assay, different amounts of recombinant N protein were spiked into PBS buffer and tested by the Microbubbling SARS-CoV-2 Antigen Assay. As shown in Figure 2A, the number of microbubbles increased linearly with the concentration of N protein between 0-20 pg/mL, with the LOD for recombinant N protein at 0.5 pg/mL (10.6 fM, blank + 3 std, n=10).

**Figure 2.**
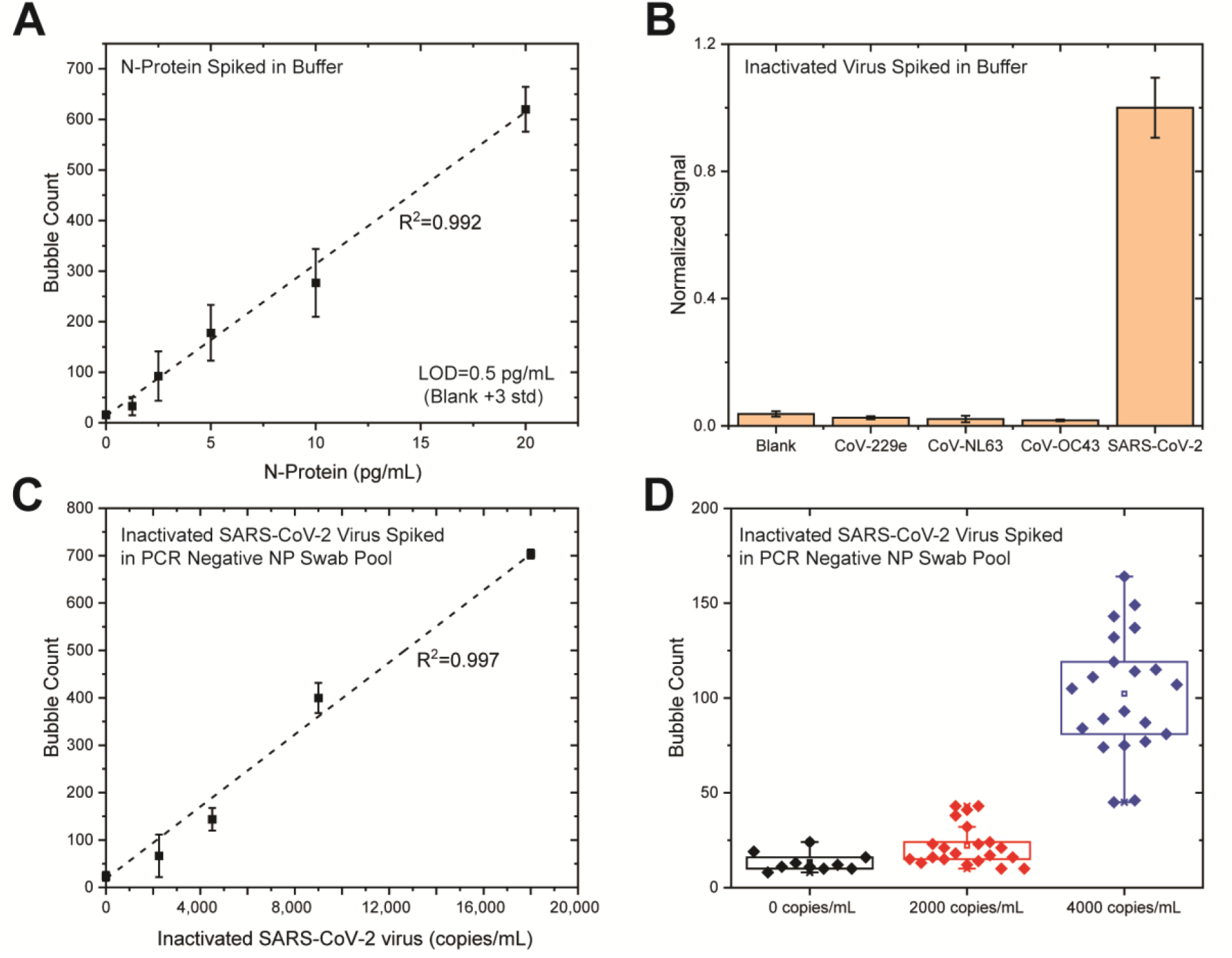
Analytical performance of the Microbubbling SARS-CoV-2 Antigen Assay. **A**. Dose response curve for spiked recombinant N protein in buffer (PBS, 1% BSA (Bovine serum albumin), pH7.4). Mean ± standard deviation; n=10 at 0, n=3 for other datapoints. **B**. Specificity of the Microbubbling SARS-CoV-2 Antigen Assay. Inactivated SARS-CoV-2 viruses were tested at 1×10^5^ copies/mL (1.4×10^4^ pfu/mL). CoV-229e, CoV-NL63 and CoV-OC43 were tested at 1×10^5^ pfu/mL. Mean ± standard deviation; n=3. **C**. Dose response curve for spiked inactivated SARS-CoV-2 viruses in rRT-PCR-negative NP swab pool. Mean ± standard deviation; n=10 at 0, n=3 for other datapoints. **D**. Determining limit of detection (LOD) of spiked inactivated SARS-CoV-2 viruses in PCR-negative NP swab pool, following the FDA antigen template guideline^11^. Assay signal (number of microbubbles) comparison between rRT-PCR-negative NP swab pool (n=10), 2000 copies/mL (n=21) and 4000 copies/mL (n=21) SARS-CoV-2 spiked into negative NP swab pool. Each diamond represents one independent experiment.

To assess the analytical specificity of the Microbubbling SARS-CoV-2 Antigen Assay towards different strains of coronaviruses, we challenged the assay with 3 human coronaviruses: CoV-229e, CoV-NL63 and CoV-OC43. Human coronaviruses OC43, 229E and NL63 cause common cold and bronchiolitis, and are the pathogens that are likely to be present in respiratory samples and possibly cross react in the Microbubbling SARS-CoV-2 Antigen Assay based on sequence homology. As shown in Figure 2B, no significant difference in signal was observed between the blank and the 3 coronaviruses (1×10^5^ pfu/mL), while over 10 times signal increase was observed for SARS-CoV-2 (1×10^5^ copies/mL or 1.4×10^4^ pfu/mL), indicating the high analytical specificity of the assay towards SARS-CoV-2.

We then evaluated the analytical performance of the assay in swab samples. Mucosal swab samples contain catalase and peroxidase, which may interfere with the Microbubbling Digital Assay by degrading the signaling reagent, H_2_O_2_. To assess this possibility, recombinant N protein was spiked into a negative pool of NP swab samples (confirmed by rRT-PCR to be negative for SARS-CoV-2) and tested in the Microbubbling SARS-CoV-2 Antigen Assay with and without PtNPs as catalyzing reagent. As shown in Supplementary Figure 2, bubbles were only observed when PtNPs were present, indicating catalase and peroxidase from the mucosal matrix had been eliminated during washing steps, and did not interfere in the Microbubbling SARS-CoV-2 Antigen Assay.

We subsequently challenged the assay with 10 rRT-PCR-negative NP swab samples. As shown in Supplementary Figure 3A, the background signals of the 10 samples varied. To investigate if the background signal was specific to the Microbubbling SARS-CoV-2 Antigen Assay, a representative sample (sample 3, with a medium background signal) was tested in an enzyme-linked immunosorbent assay (ELISA) using 96 well plate bottom to immobilize the N1 capture antibody and luciferase as the reporter enzyme labeled onto the Np detection antibody. As shown in Supplementary Figure 3B, a medium background signal was also observed for sample 3, but not for buffer control, in the ELISA assay. This indicates that the background was assay format-agnostic and intrinsic to the swab matrix. Mucins have been reported as the major cause of nonspecific bindings and background signals in many immunoassays using mucosal samples,^8^ due to their ability to bind to a variety of solid surfaces.^9,10^ Mucins are a family of glycoproteins that are widely present in the upper respiratory system^10^, with high molecular weight, high glycosylation, and abundance of negative charge groups. We postulated that mucins were the source of the background signal in our assay. To eliminate the background signal, we took advantage of the high molecular weight of mucins and used a filter with molecular weight cutoff between mucins (200 kDa∼200 MDa) and N protein (47.08 kDa) (Pierce™ protein concentrator PES, 100K MWCO) to remove mucins and retain filtrate for testing. As shown in Supplementary Figure 3C, background signals in the rRT-PCR-negative NP swab pool were removed by filtering the sample through the above concentrator, while specific signals from SARS-CoV-2 were retained.

We compared the analytical sensitivity of the assay for inactivated SARS-CoV-2 spiked in buffer (without filtration) vs. in negative NP swab pool (with filtration). As shown in Supplementary Figure 4, the analytical sensitivity in buffer (without filtration) was about 10 times higher than that in negative NP swab pool. These findings suggest that sample matrix and sample processing have significant impact on the analytical performance of the assay.

To determine the analytical sensitivity of the Microbubbling SARS-CoV-2 Antigen Assay in NP swabs, different amounts of inactivated SARS-CoV-2 viruses were spiked into an rRT-PCR-negative NP swab pool, lysed, filtered and tested using the Microbubbling SARS-CoV-2 Antigen Assay. As shown in Figure 2C, the number of microbubbles increased linearly with the concentrations of inactivated SARS-CoV-2 viruses, between 0-18000 copies/mL (virus genome RNA concentration, determined by RT-qPCR using a primer set specific to nsp14), demonstrating the quantitative capability of the assay. To determine the LOD following the FDA antigen assay template^11^, we tested 10 rRT-PCR-negative NP swab pools, 21 swab pools spiked with inactivated SARS-CoV-2 viruses at 2000 copies/mL and 21 pools at 4000 copies/mL. As shown in Figure 2D, at the concentration of 2000 copies/mL, the signals of 17 out of the 21 samples were above the mean of blank signal; at the concentration of 4000 copies/mL, the signals of all 21 samples were above the blank mean + 3std. Therefore 4000 copies/mL was determined as the LOD of the Microbubbling SARS-CoV-2 Antigen Assay for SARS-CoV-2 in swabs. This LOD translates to 400 virus copies/reaction (100 μL sample volume) in the assay.

### Clinical Performance of the Microbubbling SARS-CoV-2 Antigen Assay

We tested deidentified residual clinical NP swab samples (at least 50 samples in each category) using the Microbubbling SARS-CoV-2 Antigen Assay, and compared the results to clinical testing results using FDA EUA-approved rRT-PCR methods (Table 1, list of rRT-PCR assays in Methods). Characteristics of the clinical samples, including presence of symptoms, days after symptom onset and clinical locations were also listed in Table 1. Samples with bubble count equal to or above the count generated at LOD were determined to be positive, and those with bubble count below the LOD were determined to be negative. Compared to rRT-PCR, the positive percent agreement (PPA) was 97% (95% CI, 92-99%)in symptomatic individuals within 7 days of symptom onset and positive nucleic acid results (n=128), and the negative percent agreement (NPA) was 97% (95% CI, 94-100%) in symptomatic and asymptomatic individuals with negative nucleic acid results (n=73). The percent of antigen-positive samples decreased in individuals at 7-12 days and >12 days after symptom onset or initial COVID diagnosis, despite positive rRT-PCR results from the same samples.

**Table 1.**
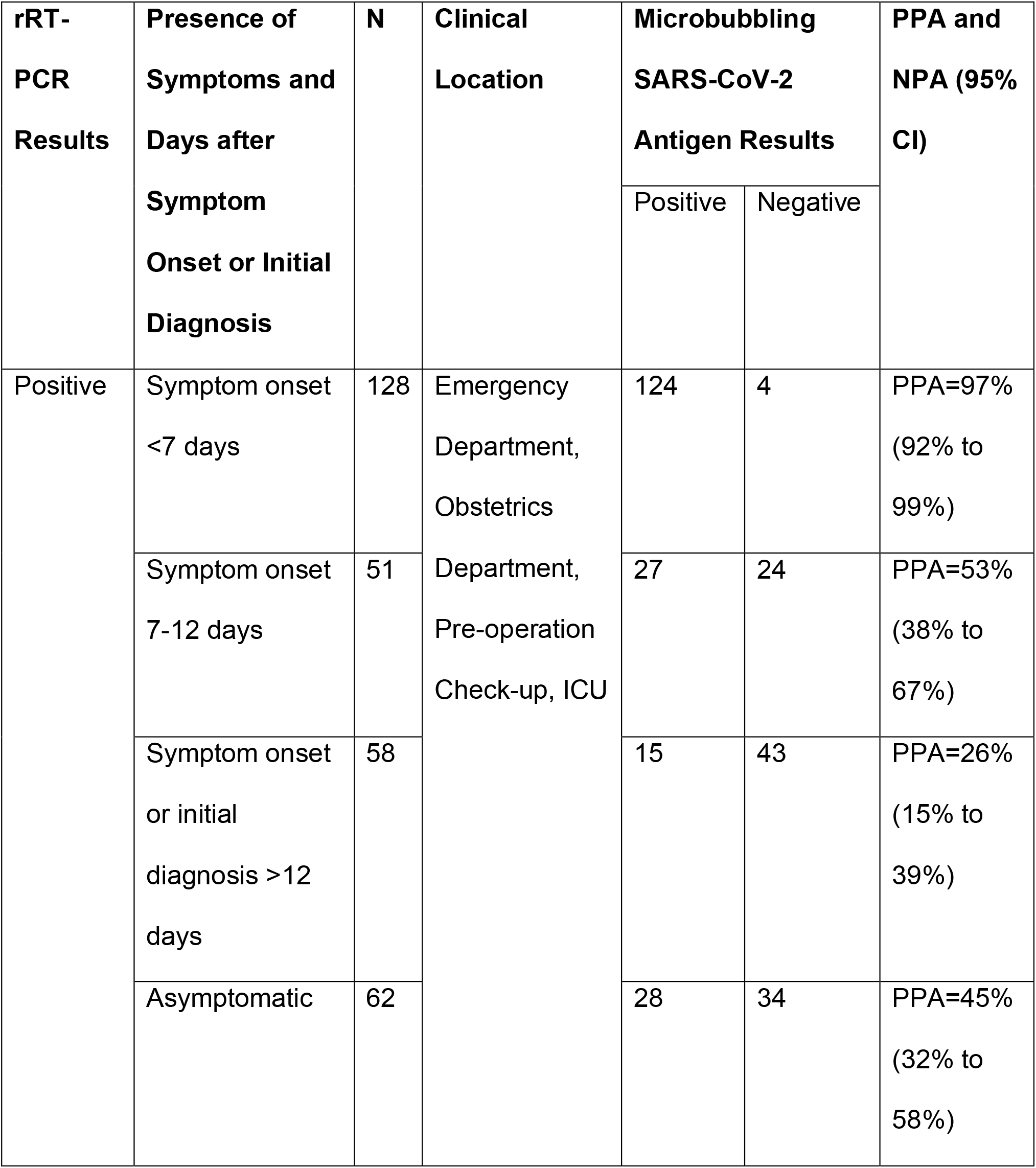
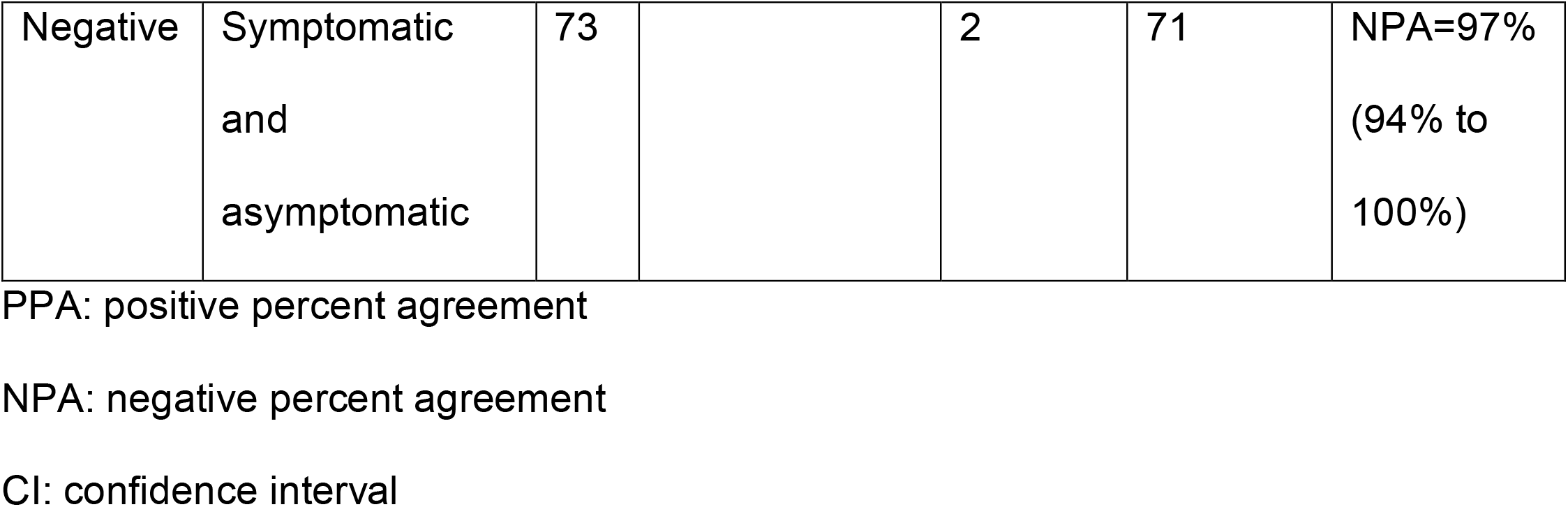
Clinical Performance of the Microbubbling SARS-CoV-2 Antigen Assay.

In the asymptomatic but rRT-PCR-positive group (n=62), the PPA was 45% (95% CI, 32-58%). The low PPA is likely due to the heterogeneous nature of this group, in which individuals may present for testing at various stages of their infections (SARS-CoV-2 testing was conducted as part of pre-surgery evaluation, prenatal care, or ED visits with non-respiratory complaints), sometimes late enough that antigens have been cleared by immune system (as we have observed in the symptomatic group). In this asymptomatic group, the Microbubbling SARS-CoV-2 Antigen Assay was able to detect a pre-symptomatic case, in which both antigen and nucleic acid results were positive in the swab sample collected one day before the patient developed cough and other respiratory symptoms. This suggests the sensitivity of the Microbubbling SARS-CoV-2 Antigen Assay is sufficiently high to detect the low concentration of antigen in pre-symptomatic cases, and the discrepancy between antigen and nucleic acid results in late samples are unlikely due to insufficient sensitivity of the Microbubbling Antigen Assay.

### Automatic Microbubble Detection Using Computer Vision and Image Classification using ML Algorithm

We have previously demonstrated that the Microbubbling Digital Assay is a platform that quantitates very low concentrations of biomarkers, with the number of microbubbles correlating linearly with the biomarker concentration in the low concentration range ^7^. In this study, we also observed that some clinical samples with low Ct values or high viral loads generated large bubbles at the time of image capture (9 min for all experiments). Therefore, we developed a computer vision algorithm to detect and locate microbubbles of any size in each image (Figure 3A) by identifying circle-like shapes. Details of this algorithm are presented in Supplementary Materials.

**Figure 3.**
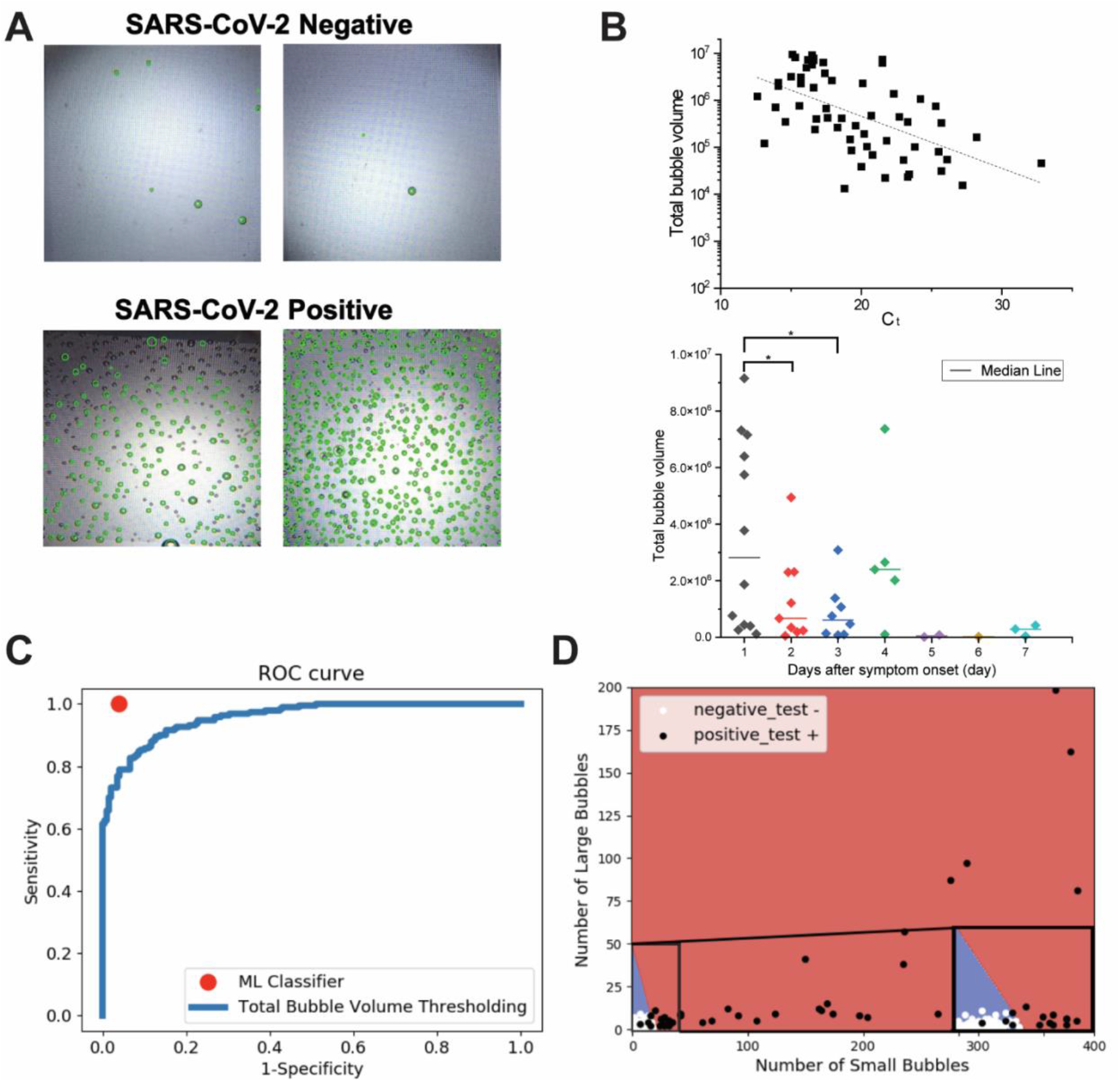
Automatic microbubble recognition, quantitation and classification using computer vision and ML algorithms. **A**. Example images from clinical NP swab samples, and microbubble detection using the computer vision algorithm. Green circles show microbubble detections from the computer vision system overlaid on the original images. **B**. Upper panel: Log transformed total bubble volume correlated inversely with Ct. Pearson linear correlation (dotted line) r= -0.56. Lower panel: Total bubble volume decreased with days-after-symptom-onset. P= 0.04 between day1 and 2, p= 0.02 between day1 and day 3 using Student’s t test. **C**. ROC curve comparing classification performance of the ML algorithm against the total bubble volume thresholding. **D**. Decision boundaries from the ML algorithm that learns linear boundaries to accurately classify images as negatives or positives in the validation data set (n=168), based on the number of automatically detected small and large microbubbles in each image. Bubbles were categorized as small if their radius was less than a heuristically set threshold of 8 pixels (in 450 × 450 pixel images, corresponding to about 50 microns), and as large if otherwise. An inset within the plot shows the enlarged area near the decision boundary.

Having automatically detected microbubble locations and sizes we add the volumes of all bubbles to estimate the total bubble volume as a quantitative readout. As shown in Figure 3B, log-transformed total bubble volume correlated inversely with Ct values, and total bubble volume decreased with days-after-symptom-onset. This indicates that total bubble volume may be a potential marker for antigen burden.

Next, we compared two approaches for automatic classification of microbubble images as positives or negatives, both based on the computer vision-based microbubble detection. First, we varied the threshold on the estimated total bubble volume to generate an ROC curve (Figure 3C), using human visual inspection-based classification as gold standard. We compared this against a ML-based classifier, built as follows. In each image, we counted the number of detected bubbles whose radius was above and below a set threshold (∼50 microns, corresponding to 8 pixels in the images). These big bubble and small bubble counts formed the 2-dimensional feature representation of each image. In this 2-dimensional space, we trained a linear separator using the linear support vector ML algorithm^14^. We randomly sampled 168 images to use as the training dataset and evaluated the performance (of both approaches) on the remaining 168 images (87 negatives, and 81 positives). Fig 3C shows both the ROC curve of the total bubble volume-based classifier, as well as the sensitivity and specificity of the 2D bubble count features-based ML classifier. The ML classifier performs with much higher sensitivity (100%) and specificity (95%) than any point on the bubble volume thresholding ROC curve. The decision boundaries identified by the ML classifier is shown in Figure 3D. These results were very stable across three random 50-50 splits of the full dataset (336 images) into training and testing data. Overall, this demonstrates that fully automated image analysis and ML are able to produce antigen burden quantitation and accurate classification from the Microbubbling SARS-CoV-2 Antigen assay. For easy reproducibility, we will publicly release all code for computer vision-based microbubble detection and ML.

### Tracking Antigen Dynamics in Different Patient Populations

In the clinical validation group (Table 1), antigen and nucleic acid results showed limited correlation in individuals after the acute infection phase (>7 days). To further investigate this phenomenon longitudinally, we tested for antigen using the Microbubbling SARS-CoV-2 Antigen Assay in serial NP/oropharyngeal (OP) swabs collected from 38 ICU patients hospitalized due to COVID (Figure 4A). To compare nucleic acid with antigen dynamics, N1 gene copy number quantitated using RT-qPCR were also plotted in Figure 4A. Consistent with what was observed in Table 1, nucleic acid and antigen results did not always correlate with each other in this patient cohort. Many serial samples did not have detectable N antigens despite significant copy numbers of the N1 gene (>>4000 copies/mL). Several patients (eg. #406, #391, #257) converted from N antigen positive to antigen negative at various time points despite that N1 gene copies either increased or remained stable and above LOD of the Microbubbling assay. This suggests the RT-qPCR may be detecting N1 gene fragments that were not producing intact N protein with the epitope for antibody recognition. On the other hand, for patient #401, N1 gene copy number increase correlated with the change of N antigen result from negative to positive, possibly indicating active virus replication during this period.

**Figure 4.**
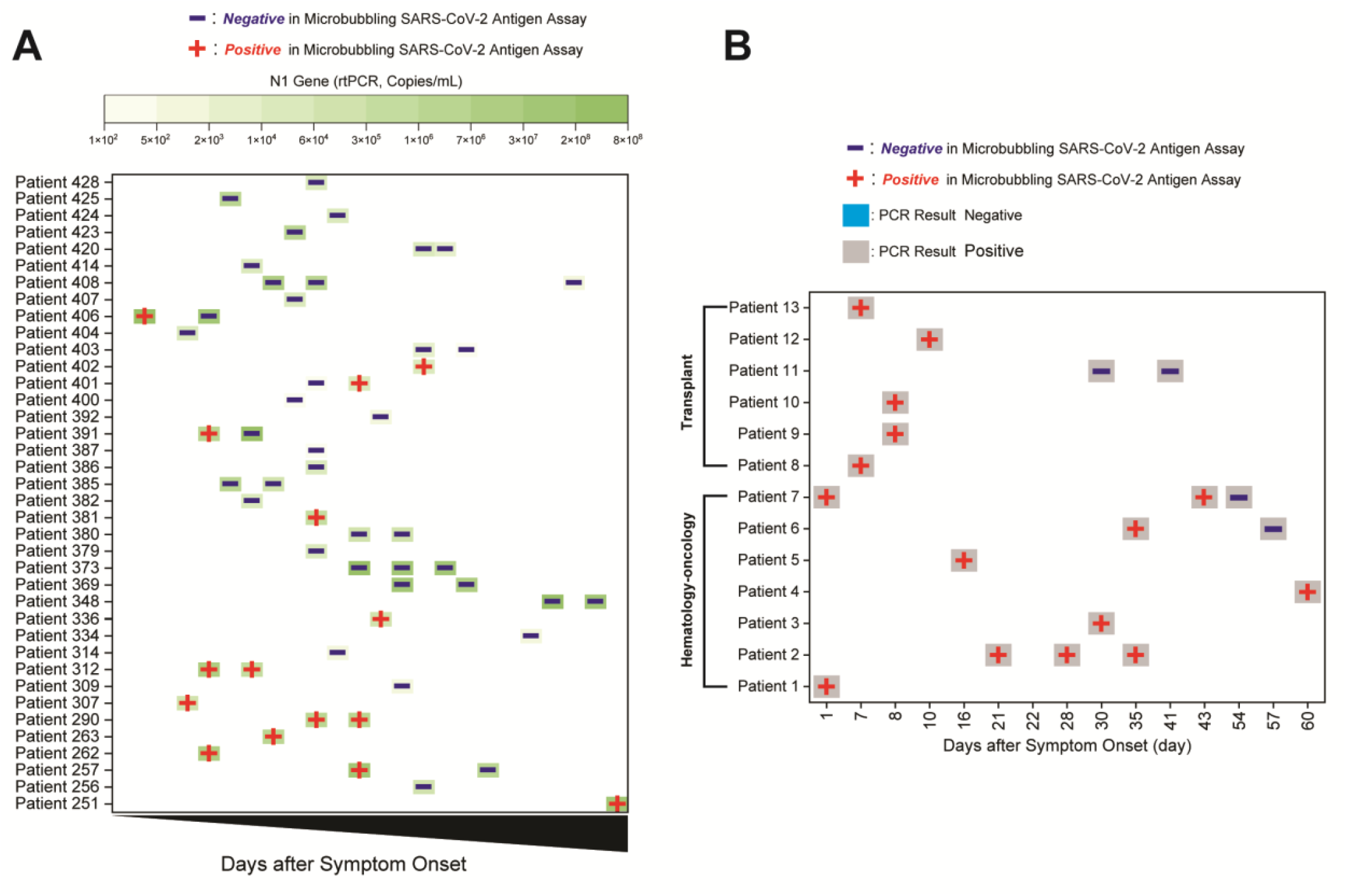
Tracking antigen dynamics using the Microbubbling SARS-CoV-2 Antigen Assay in serial swab samples in **A**. COVID inpatients in ICU and **B**. immunocompromised COVID patients with either hematological malignancies or transplants. A red + indicates a positive antigen result, whereas a blue - indicates a negative antigen result. Color intensities of the green squares in A. indicate the quantitative results of N1 gene copy number using RT-qPCR. Colors of the squares in B. indicate the qualitative nucleic acid results using FDA EUA-approved rRT-PCR methods.

Immunocompromised individuals remained antigen positive for a longer period of time in our validation group (Table 1). Out of 4 immunocompromised and nucleic acid-positive patients, who either had hematological malignancies or transplants, and were 7-12 days after symptom onset, all 4 were antigen positive (100% positivity rate, compared to 53% in the entire validation group in the 7-12 days range). Out of 13 immunocompromised and nucleic acid-positive patients >12 days after symptom onset, 9 remained antigen positive (69% positivity rate, compared to 26% in the entire validation group in the >12 days range). It has been reported that immunocompromised individuals with hematological malignancies have prolonged viral clearance and delayed or absent humoral responses^12^. We therefore tracked antigen results at various days-after-symptom-onset in a group of 13 immunocompromised patients with either hematological malignancies or transplants (Figure 4B). Notably, nucleic acid was detected in all samples for these patients, while antigen was detectable for various periods of time. For the 6 patients for whom we had samples at or after 30 days after symptom onset, 5 remained antigen positive around 30 days. Two patients (#7 and #6) converted from antigen positive to negative at 54 and 57 days, respectively. The furthest antigen-positive timepoint observed in this group was from a patient (#4) post CAR-T therapy for diffuse large B-cell lymphoma, who continued to have respiratory issues and tested positive for both nucleic acid and antigen 2 months after initial COVID diagnosis. Our results indicate that immunocompromised individuals have delayed and variable antigen clearance timeline, which correlates poorly with their nucleic acid results.

## Discussion

We have demonstrated that the Microbubbling SARS-CoV-2 Antigen Assay can detect the N antigen with an analytical sensitivity of 0.5 pg/mL (10.6 fM) and high specificity. The LOD in buffer is 100 times better than commercially available SARS-CoV-2 N protein ELISA kits (Supplemental Table 1). The LOD in swabs (4000 copies/mL) is comparable to many FDA EUA-approved rRT-PCR assays (10^2^-10^5^ copies/mL), better than current EUA-approved lateral flow antigen tests, the majority of emerging SARS-CoV-2 antigen assays and CRISPR-based tests (Supplemental Table 1). Clinical validation comparing to EUA-approved rRT-PCR methods showed excellent PPA and NPA in both symptomatic and asymptomatic individuals for acute infection detection (<7 days). Residual NP swabs in 3mL saline tubes were used for convenience during our clinical validation. We expect higher PPA using swabs directly reconstituted using a lower volume of extraction reagents, which can be tested in future studies. These findings suggest that the Microbubbling SARS-CoV-2 Antigen Assay may be a valuable tool in detecting acute SARS-CoV-2 infections. The gradual decrease in PPA in samples after 7 days of symptom onset is consistent with seroconversion^13^ and clearance of antigen from the body. It has been suggested that infectivity of individuals with high Ct values and symptom onset to testing > 8 days is low^14^, and it was hypothesized that the sensitive molecular methods detect viral genomic fragments, rather than transmissible viruses ^4,15,16^. Our data in Table 1 and Figure 4A are consistent with this hypothesis. The fact that the Microbubbling Antigen Assay was able to detect a pre-symptomatic infection case indicates that the assay has the required sensitivity to detect pre-symptomatic antigen levels. The lower PPA in asymptomatic individuals is likely due to the heterogeneity of the asymptomatic group, some of whom might have presented to the study after antigen had been cleared by the immune system while genomic fragments remained.

We have also demonstrated that total bubble volume, a quantitative output from the Microbubbling SARS-CoV-2 Antigen Assay, correlated inversely with Ct values and days-after-symptom-onset, and may serve as a potential marker for antigen burden. This allows us to probe and monitor the level and dynamics of virus antigens at different stages of infection, and to explore the correlation between antigen burden and disease severity/prognosis in future studies, which may be valuable in advancing understanding of the SARS-CoV-2 virus. With the exception of some reports focusing on antigens in blood ^17,18^ ,very little is known about the burden and dynamics of SARS-CoV-2 antigens in respiratory samples and other body fluids during the infection course. Although it is widely reported that infected individuals may test positive for viral nucleic acids for weeks or months despite symptom resolution, it is unknown whether viral antigens also remain positive for extended time in these individuals. Our data demonstrated that in immunocompetent individuals, antigen is often cleared much faster than nucleic acid in respiratory samples. On the other hand, we also showed that immunocompromised individuals remained antigen positive for much longer periods of time, and cleared antigen at various timepoints. In general, antigen dynamics did not correlate well with that of nucleic acids outside of the acute infection window. Simultaneous positivity for both antigen and nucleic acid in the same respiratory sample may be an indicator of actively replicating viruses, and necessity for virus genome sequencing to monitor for variants in prolonged infections, and may also inform the appropriate allocation and utilization of isolation resources (negative pressure rooms, PPE etc.) for patient management.

We expect that the Microbubbling Antigen Assay will be able to detect acute infections earlier than lateral flow assays, thereby mitigating concerns for false negatives using rapid antigen tests. However, we did not test this hypothesis in a side-by-side comparison study. Another limitation of the study is that the data of days-since-symptom-onset came from limited documentation in the medical records based on patient self-reporting. These data may not always be accurate or complete. Ct values were not available from some rRT-PCR methods used in this study, and are in general not well-standardized across platforms ^19^. This may lead to heterogeneity in the nucleic acid data presented. We had access to a relatively small number of serial samples from immunocompromised patients. Finally, due to the poor culturability of most clinical specimens, we did not use viral culture to assess the infectivity of antigen positive samples.

In future studies, the Microbubbling SARS-CoV-2 Antigen Assay can be automated and multiplexed with other infectious disease antigens, and applied to other sample types. With the convenient computer vision and ML based algorithms on smartphone for automated bubble recognition, quantitation and result classification, this assay also has the potential to be used at the point-of-care for frequent and repeated testing. Our relatively simple computer vision and ML pipeline produced accurate automatic classification results, which we expect will be further improved through training more sophisticated deep learning-based systems on larger datasets. In particular, a common failure mode in our current visual bubble detection system is when bubbles appear blurry because they are not on the focal plane, or are inconsistently lit in different portions of an image. These may be overcome through a bubble detector that, like the ML classifier, is also learned from data, rather than the current system that is engineered to locate circle-like shapes in the image. Finally, in an integrated system, the microbubble results could also be conveniently uploaded into a database that integrates other clinical parameters and molecular/serology results etc. to provide a comprehensive picture of the disease in the population, enabling disease tracking and future predictive algorithm development.

In conclusion, we have demonstrated that the Microbubbling Digital Antigen Assay can be applied to the detection of SARS-CoV-2 antigen with high sensitivity and specificity. The Microbubbling SARS-CoV-2 Antigen Assay demonstrated high PPA and NPA with rRT-PCR methods in symptomatic and asymptomatic individuals early in the infection course. We have developed computer vision and ML algorithms to quantitate and classify the image output, and also shown that the assay can be a valuable tool in providing insights into antigen dynamics in various patient populations.

## Data Availability

All data referred to in the manuscript is available from the research team. The OpenCV-based Python code for the computer vision and machine learning pipeline is available at the following address: https://github.com/jianingq/microbuble-detection-and-classification.git.

## Supplementary Materials

Materials and Methods, Supplemental Figures and Table are available online.

## Acknowledgement

HC, ZL and PW have received support from National Institute of Health grants R01DA035868, R01EB029363 and National Science Foundation grant 1928334. SRW has received support from National Institute of Health grant R01AI40442 and Penn Center for Research on Coronaviruses and Other Emerging Pathogens. We thank the RADx-Tech Program, Penn Center for Precision Medicine, Penn Health-Tech and Penn Center for Innovation & Precision Dentistry for providing funding for this project.

This work was carried out in part at the Singh Center for Nanotechnology, part of the National Nanotechnology Coordinated Infrastructure Program, which is supported by the National Science Foundation grant NNCI-2025608.

## References

1 Wang, X. et al. Limits of Detection of 6 Approved RT-PCR Kits for the Novel SARS-Coronavirus-2 (SARS-CoV-2). Clin Chem 66, 977–979, doi:10.1093/clinchem/hvaa099 (2020).

2 Shi, J., Han, D., Zhang, R., Li, J. & Zhang, R. Molecular and Serological Assays for SARS-CoV-2: Insights from Genome and Clinical Characteristics. Clin Chem 66, 1030–1046, doi:10.1093/clinchem/hvaa122 (2020).

3 SARS-CoV-2 Reference Panel Comparative Data. https://www.fda.gov/medical-devices/coronavirus-covid-19-and-medical-devices/sars-cov-2-reference-panel-comparative-data

4 Henderson, D. K. et al. The perplexing problem of persistently PCR-positive personnel. Infect Control Hosp Epidemiol, 1–2, doi:10.1017/ice.2020.343 (2020).

5 Coronavirus (COVID-19) Update: FDA Authorizes First Antigen Test to Help in the Rapid Detection of the Virus that Causes COVID-19 in Patients., <https://www.fda.gov/news-events/press-announcements/coronavirus-covid-19-update-fda-authorizes-first-antigen-test-help-rapid-detection-virus-causes> (2020).

6 Diao, B. et al. Accuracy of a nucleocapsid protein antigen rapid test in the diagnosis of SARS-CoV-2 infection. Clinical microbiology and infection : the official publication of the European Society of Clinical Microbiology and Infectious Diseases 27, 289.e281-289.e284, doi:10.1016/j.cmi.2020.09.057 (2021).

7 Chen, H. et al. Quantitation of Femtomolar-Level Protein Biomarkers Using a Simple Microbubbling Digital Assay and Bright-Field Smartphone Imaging. Angewandte Chemie 131, 14060–14066, doi:10.1002/ange.201906856 (2019).

8 Helton, K. L., Nelson, K. E., Fu, E. & Yager, P. Conditioning saliva for use in a microfluidic biosensor. Lab Chip 8, 1847–1851, doi:10.1039/b811150b (2008).

9 McColl, J., Yakubov, G. E. & Ramsden, J. J. Complex Desorption of Mucin from Silica. Langmuir 23, 7096–7100, doi:10.1021/la0630918 (2007).

10 Song, D., Cahn, D. & Duncan, G. A. Mucin Biopolymers and Their Barrier Function at Airway Surfaces. Langmuir 36, 12773–12783, doi:10.1021/acs.langmuir.0c02410 (2020).

11 Coronavirus Disease 2019 (COVID-19) Emergency Use Authorizations for Medical Devices/ In Vitro Diagnostics EUAs, <https://www.fda.gov/medical-devices/coronavirus-disease-2019-covid-19-emergency-use-authorizations-medical-devices/vitro-diagnostics-euas> (2020).

12 Abdul-Jawad, S. et al. Acute Immune Signatures and Their Legacies in Severe Acute Respiratory Syndrome Coronavirus-2 Infected Cancer Patients. Cancer Cell 39, 257–275 e256, doi:10.1016/j.ccell.2021.01.001 (2021).

13 Huang, A. T. et al. A systematic review of antibody mediated immunity to coronaviruses: kinetics, correlates of protection, and association with severity. Nat Commun 11, 4704, doi:10.1038/s41467-020-18450-4 (2020).

14 Bullard, J. et al. Predicting infectious SARS-CoV-2 from diagnostic samples. Clin Infect Dis, doi:10.1093/cid/ciaa638 (2020).

15 Jefferson, T., Spencer, E., Brassey, J. & Heneghan, C. Viral cultures for COVID-19 infectivity assessment. Systematic review. MedRxiv, doi:10.1101/2020.08.04.20167932 (2020).

16 Rhee, C., Kanjilal, S., Baker, M. & Klompas, M. Duration of Severe Acute Respiratory Syndrome Coronavirus 2 (SARS-CoV-2) Infectivity: When Is It Safe to Discontinue Isolation? Clin Infect Dis, doi:10.1093/cid/ciaa1249 (2020).

17 Ogata, A. F. et al. Ultra-sensitive Serial Profiling of SARS-CoV-2 Antigens and Antibodies in Plasma to Understand Disease Progression in COVID-19 Patients with Severe Disease. Clin Chem, doi:10.1093/clinchem/hvaa213 (2020).

18 Hingrat, Q. L. et al. Detection of SARS-CoV-2 N-antigen in blood during acute COVID-19 provides a sensitive new marker and new testing alternatives. Clin Microbiol Infect, doi:10.1016/j.cmi.2020.11.025 (2020).

19 Rhoads, D. et al. College of American Pathologists (CAP) Microbiology Committee Perspective: Caution must be used in interpreting the Cycle Threshold (Ct) value. Clin Infect Dis, doi:10.1093/cid/ciaa1199 (2020).

