## Supplementary Materials for "Femtomolar SARS-CoV-2 Antigen Detection Using the Microbubbling Digital Assay with Smartphone Readout Enables Antigen Burden Quantitation and Dynamics Tracking"

[1]Department of Pathology and Laboratory Medicine, [2]Bioengineering Graduate Program, [3]Department of Computer and Information Science and GRASP Lab, [4]Department of Medicine, [5]Department of Microbiology and Penn Center for Research on Coronavirus and Other Emerging Pathogens, [6]Department of Orthodontics, Divisions of Pediatric Dentistry and Community of Oral Health, School of Dental Medicine, [7]Center for Innovation & Precision Dentistry, School of Dental Medicine and School of Engineering and Applied Sciences, [8]Department of Oral Medicine, School of Dental Medicine and [9] Center for Clinical and Translational Research, University of Pennsylvania, Philadelphia, PA

[a] These two authors contributed equally.

*Address correspondence to:

Ping Wang

Department of Pathology and Laboratory Medicine

University of Pennsylvania

3400 Spruce St., Founders 7.103

Philadelphia, PA

### Supplemental Materials

#### Materials and Methods

##### Materials

Materials and supplies for the Microbubbling Digital Assay were reported previously [1]. Additional materials and supplies used in this study are listed below. TWEEN® 20 (Molecular Biology Grade, P9416-100ML) was purchased from Sigma-Aldrich, Inc. (St. Louis, MO, USA). Halt™ Protease Inhibitor Cocktail (78439) and Pierce™ protein concentrators PES, 100K MWCO, 0.5 mL (88503) were purchased from Thermo Fisher Scientific, Inc. (Rockford, IL, USA). Antibodies raised against N-protein from SARS-CoV and SARS-CoV-2, and recombinant N-Protein (40588-V08B) were purchased from Sino Biological Inc. (Wayne, PA, USA).

##### Clinical Swab Samples

The study was approved by the Institutional Review Board of the University of Pennsylvania. The NP swab samples used for clinical performance validation (Table 1) and from immunocompromised individuals were collected from patients presented to the Hospital of University of Pennsylvania as part of the routine clinical care to test for SARS-CoV-2, and transported to the clinical laboratory in 3 mL saline tube (Becton Dickinson cat#15439) according to standard clinical operating procedure. These samples were tested according to the clinical protocols using either the Xpert Xpress SARS-CoV-2 or SARS-CoV-2/Flu/RSV assay (Cepheid, Sunnyvale, CA), the Simplexa COVID-19 Direct kit (DiaSorin Molecular LLC, Cypress, CA), the cobas Liat SARS-CoV-2 & Influenza A/B assay (Roche, Basel, Switzerland), or the ePlex Respiratory Pathogen Panel 2 (GenMark, Carlsbad, CA). Residual swab samples were then transferred to the research lab within 48 hrs of sample collection. Once the samples were in the research lab, Halt™ Protease inhibitor cocktail was immediately added at 1:100 v/v ratio following manufacture's instruction, and the samples were then stored at -80°C. Before the microbubbling testing, samples were thawed at room temperature. Ten percent TWEEN® 20 was added to thawed samples to reach the final concentration of 0.1%, which were immediately tested using the Microbubbling SARS-CoV-2 Antigen Assay at room temperature. Controls (inactivated SARS-CoV-2 spiked into negative NP swab pool) were processed in the same way.

The swabs from ICU patients were collected from patients hospitalized at the Hospital of University of Pennsylvania with documented COVID. Following informed consent obtained under protocol #823392 approved by the University of Pennsylvania IRB, NP and oropharyngeal (OP) swabs were obtained and eluted together in 3 mL of viral transport media (VTM, HBSS with 2% FBS, 100 I.U./mL penicillin, 100 µg/mL streptomycin, 100µg /mL Gentamicin, 0.5µg /mL Amphotericin). These samples were stored at -80°C before being thawed and tested using the Microbubbling Antigen Assay. In parallel, RNA was extracted from 140 ul of sample using the QIAamp Viral RNA Mini Kit (Qiagen, Hilden, Germany) and 3 µl RNA was subject to SARS-CoV-2 genome quantification by RT-qPCR using the CDC N1 primer set as described[2].

**Cell culture and virus titration**

Vero E6 cells (ATCC), monkey kidney epithelial cells, were maintained in DMEM supplemented with 10% (v / v) FBS, 1% (v / v) penicillin / streptomycin, 1% (v / v) L-glutamine, 10mM HEPES at 37°C and 5% CO2 incubator. SARS-CoV-2 was obtained from BEI (WA-1 strain). SARS-CoV-2 virus stocks were prepared by Vero E6 cell infections with media containing 2% FBS. After 5 days of virus infection, the cells were frozen and thawed. The media were collected and centrifuged 3,000 rpm for 20 min. Viral titer was determined by 50% tissue culture infective doses (TCID50) of Reed-Muench method[3]. In brief, 96 well plate with Vero E6 cells were infected by serial 10 fold diluted SARS-CoV-2. Two days post infection, the cells were fixed by 4% formaldehyde and analyzed infected cells by automated microscopy. The cells were stained for nuclei (Hoechst 333342) and J2 (dsRNA) antibody. At least 4 replicates were used for each set. Values corresponding to TCID50 were calculated according to the method of Reed and Muench[4].

Supernatant from SARS-CoV-2 culture was inactivated by heating at 56 °C for 1 hour, aliquoted and frozen at -80 °C. The supernatant is determined to have a virus concentration of $3.6x10^7$ genome copies/mL using a primer set specific to nsp14 and RT-qPCR, equivalent to $5x10^6$ pfu/ml and $6.3x10^5$ $TCID_{50}$/mL before inactivation. Briefly, viral RNA was purified using Trizol (Invitrogen) followed by RNA Clean and Concentrate kit (Zymo Research). For cDNA synthesis, reverse transcription was performed with random hexamers and Moloney murine leukemia virus (M-MLV) reverse transcriptase (Invitrogen). SARS-CoV-2 nsp14 primers and SYBR green master mix (Applied Biosystems) was used in a QuantStudio 6 Flex RT-PCR system (Applied Biosystems). To quantify absolute copy numbers of SARS-CoV-2, Synthetic RNA from SARS-Related Coronavirus 2 was used as a standard (BEI, NR-52358). Primer sequences are as like follow. Nsp14-F: 5'- TGGGGYTTTACRGGTAACCT -3', Nsp14-R: 5'- AACRCGCTTAACAAAGCACTC -3'.

Cultured human coronaviruses OC43, 229E and NL63 stocks were prepared under BSL 2 Laboratory conditions at University of Pennsylvania, aliquoted and frozen at -80°C. Aliquots of these virus culture samples were subsequently thawed, diluted and tested using the Microbubbling SARS-CoV-2 Antigen Assay.

**Fabrication of Microbubbling Microchips**

Microbubbling microchips were fabricated as reported previously [1]. Briefly, a cover glass as the bottom supporting layer, a polydimethylsiloxane (PDMS) sheet with an array of 10,000 micro wells as the middle layer, and a PDMS top layer containing a round sample chamber was assembled together. A 3 µm thick parylene C layer was then coated on the assembled microchip surface.

**Functionalization of Superparamagnetic Microbeads and Platinum Nanoparticles**

Superparamagnetic microbeads and platinum nanoparticles were functionalized as previously reported [1]. Briefly, LodeStars® High Bind carboxyl-terminated superparamagnetic beads were functionalized with antibody using EDC-NHS coupling, blocked with 1% BSA, and then resuspended in PBS buffer containing 1% BSA and 0.02% sodium azide. NeutrAvidin was conjugated to the surface of platinum nanoparticles (PtNPs) through sulfo-Pt bonds by mixing in citrate buffer overnight at 4 °C, blocked with 1% BSA, and then resuspended in PBS buffer containing 1% BSA.

**Microbubbling SARS-CoV-2 Antigen Assay**

Clinical NP swab eluant samples (200 µL) were first mixed with virus lysis buffer of 10% Tween20 (100x, 2 µL) and 100x protease inhibitor cocktail (2 µL) and incubated at room temperature for 30 min. The lysed sample was then processed using the centrifugal filter (Pierce™ protein concentrators PES, 100K MWCO) by centrifuging at 12000g for 10min collecting the filtrate for following assays. Similar protocol for the Microbubbling Digital Assay was then followed as previously published [1], with the exception that the incubation time lengths were 30 min each, which were optimized to reduce assay time without sacrificing performance. Briefly, sample solutions (100 µL) were incubated with suspensions of 500,000 capture antibody functionalized magnetic beads, on a roller (12 rpm) at room temperature for 30 min. The beads were then separated using magnets and washed 3 times with PBS buffer pH 7.4 containing 0.01% TWEEN® 20, and then resuspended in 100 µL of 250 ng/mL biotinylated detection antibody in PBS containing 1% BSA, on a roller (12 rpm) at room temperature for 30 min. The beads were then separated using magnets and washed 3 times with PBS buffer pH 7.4 containing 0.01% TWEEN® 20, and then resuspended in 100 µL of 1 µg/mL NeutrAvidin functionalized PtNP in PBS containing 1% BSA, on a roller (12 rpm) at room temperature for 30 min. The beads were then separated using magnets and washed 3 times with PBS buffer pH 7.4 containing 0.01% TWEEN® 20 and resuspended in 100 µL of 30% $H_2O_2$. The magnetic beads slurries were then added into the chambers of the microbubbling microchips. The microbubbling microchips were placed on neodymium disc magnets for 1 min to pull down the beads to the bottom of the microchips. Images of microbubbles in the microwell arrays were captured with a smart phone camera with a mobile microscope attachment.

##### Imaging and Analysis of Microbubbling Assay Output

Microbubbles on the microbubbling microchips were imaged using an iPhone 11 or an iPad with the uHandy mobilephone microscope (9x, 5 mm focusing length,

Aidmics Biotechnology Co. Taipei, Taiwan). A computer vision system was designed for automatically processing images to identify, count and measure microbubbles and calculate Total Bubble Volume. The system detects and locates microbubbles through two parallel approaches before merging the detections. The first approach employs Canny edge detection [5] to find microbubble contours, then computes the minimum bounding circle around each detected microbubble to identify its location and size. The second approach applies the Hough Circle transform [6] to improve detection of larger bubbles. Results from the two approaches were then merged. To avoid double counting bubbles detected through both approaches, those bubbles detected by the second approach, that have large overlap with the bubbles detected by the first approach were removed. The OpenCV[7]-based Python code for the computer vision and machine learning pipeline is available at the following address: https://github.com/jianingq/microbuble-detection-and-classification.git.

**Limit of Detection (LOD) Determination**

Recombinant N antigen was diluted to different concentrations and tested using the Microbubbling SARS-CoV-2 Antigen Assay. The number of bubbles generated was plotted against the N antigen concentration. The LOD for recombinant N antigen was determined as blank + 3 standard deviation (SD) of the blank (10 datapoints for blank).

To determine the LOD for inactivated SARS-CoV-2 in negative NP swab pool, the method defined in the FDA SARS-CoV-2 Antigen Template for Test Developers was used [8]. Inactivated supernatant from SARS-CoV-2 culture was serially diluted to different concentrations in negative NP swab pool, processed using the Pierce™ 100K MWCO centrifugal filter and tested using the Microbubbling SARS-CoV-2 Antigen

Assay. A negative pool and three serial virus dilutions were first tested in triplicate to identify the lowest concentration with three positive readouts. Twenty-one replicates of that concentration were then repeated. The lowest concentration at which at least 19 of the 20 replicates were positive was determined to be the LOD.

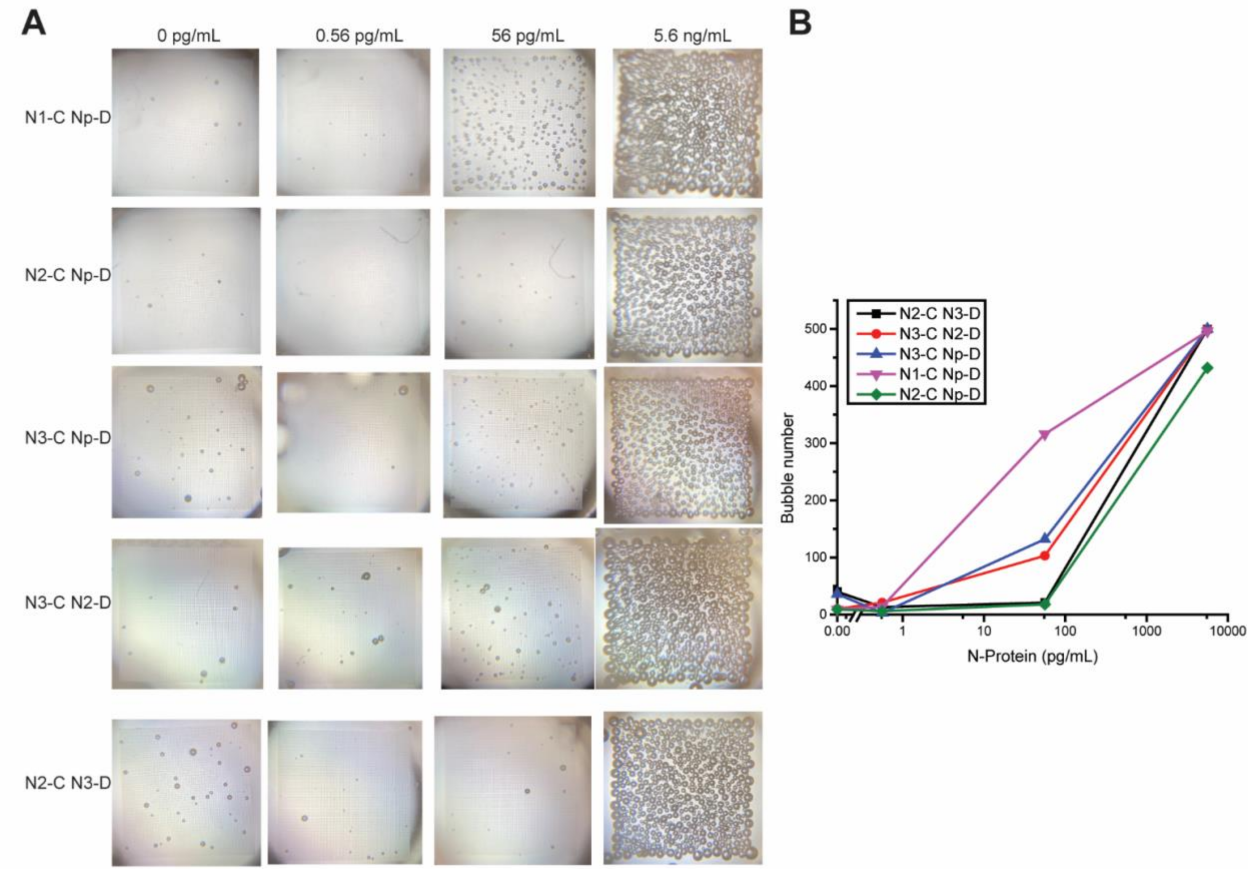


**Supplementary Figure. 1**. Screening of the antibody pairs and capture/detection combinations for the detection of SARS-CoV-2 N antigen using Microbubbling Assay **A.** Microbubble images at different concentrations of SARS-CoV-2 N antigen with antibody pairs and capture/detection combinations. N1-C Np-D: N1 as capture antibody, Np as detection antibody; N2-C Np-D: N2 as capture antibody, Np as detection antibody; N3-C Np-D: N3 as capture antibody, Np as detection antibody; N3-C N2-D: N3 as capture antibody, N2 as detection antibody; N2-C N3-D: N2 as capture antibody, N3 as detection antibody. **B**. Dose response curves with antibody pairs and capture/detection combinations.

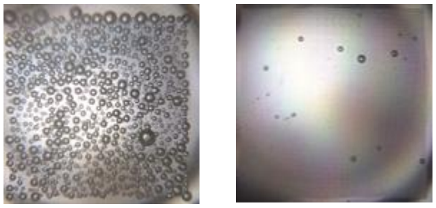


**Supplementary Figure. 2**. Catalase and peroxidase in mucosal samples do not interfere in the Microbubbling SARS-CoV-2 Antigen Assay.

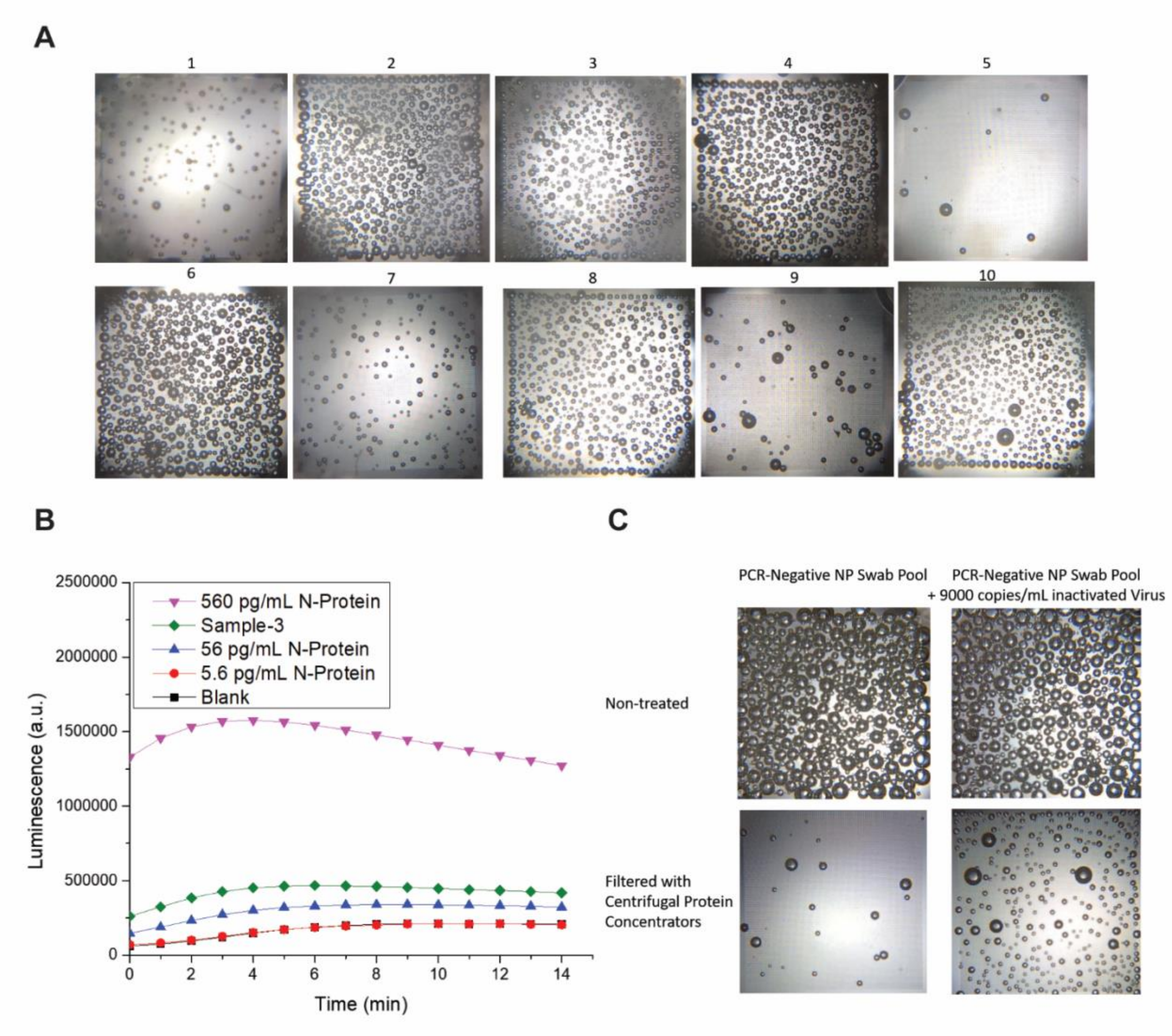


**Supplementary Figure. 3**. Study of the background signals in nasopharyngeal (NP) swab samples. A) Varied background signals in 10 rRT-PCR negative NP swab samples. B) Background signal of rRT-PCR negative NP swab sample #3 in a luminescence-based ELISA. C) Background signal was removed in an rRT-PCR negative NP swab pool by filtering using the centrifugal protein concentrator (Pierce™ protein concentrators PES, 100K MWCO, 0.5 mL, 88503).

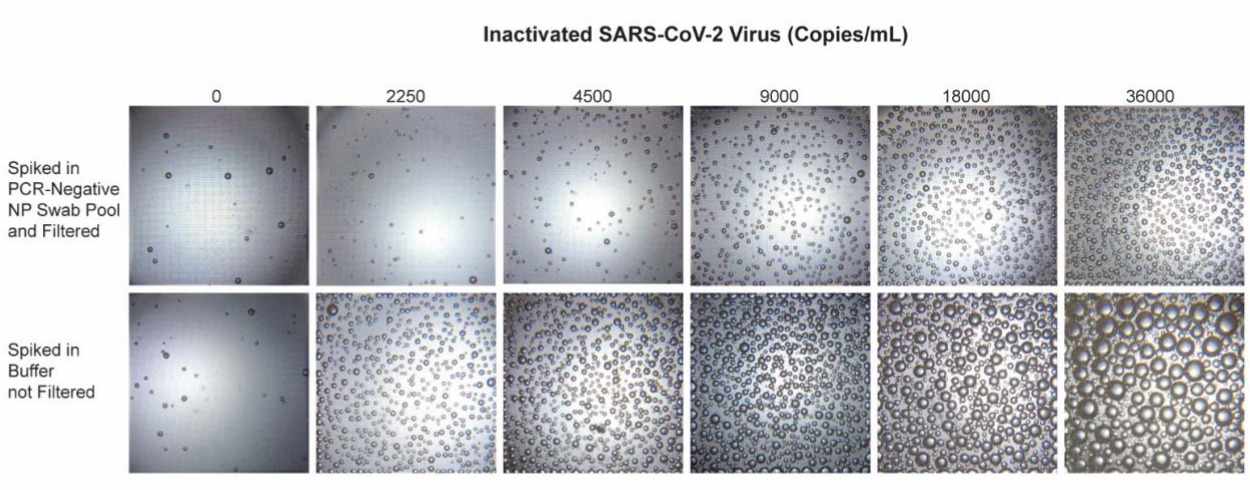


**Supplementary Figure. 4**. Compare the analytical performance of the Microbubbling SARS-CoV-2 Antigen Assay for the detection of inactivated SARS-CoV-2 virus in buffer vs. in negative NP swab pool. Microbubble images at different virus concentrations, in negative NP swab pool and filtered (upper row) vs. buffer and not filtered (lower row).

**Supplementary Table 1.** Summary of the performance of currently available and emerging assays for SARS-CoV-2 detection.

| Assay name | | Method | Target | LOD | Performance in Clinical Samples | Ref. |
|---|---|---|---|---|---|---|
| **EUA Approved SARS-CoV-2 Antigen Tests** | CareStart ™ COVID-19 Antigen | LFA | Nucleocapsid protein antigen | 8 x $10^2$ $TCID_{50}$/ml | Nasopharyngeal swab samples 5 days since symptom onset Positive Percent Agreement (PPA) 87.18% | 9 |
| | LumiraDx SARS-CoV-2 Ag Test | Microfluidic Immunofluorescence Assay | Nucleocapsid protein antigen | 32 $TCID_{50}$/mL | Nasal Swab Samples 12 days since symptom onset PPA 97.6% | 10 |
| | BD Veritor System for Rapid Detection of SARS-CoV-2 | Chromatographic Digital Immunoassay | Nucleocapsid protein antigen | 1.4 x $10^2$ $TCID_{50}$/mL | Nasal swab samples 5 days since symptom onset PPA 84% | 11 |
| | VITROS Immunodiagnostic Products SARS-CoV-2 Antigen Reagent Pack | Chemiluminescence Immunoassay | Nucleocapsid protein antigen | From 5.0 x $10^2$ $TCID_{50}$/mL to 1.5 x $10^3$ $TCID_{50}$/mL. | Nasopharyngeal swab samples 5 days since symptom onset PPA 80% | 12 |
| | Simoa SARS-CoV-2 N Protein Antigen Test | Paramagnetic Microbead-based Immunoassay | Nucleocapsid protein antigen | 0.31 $TCID_{50}$/mL | Nasopharyngeal swab samples 7 days since symptom onset PPA 100% | 13 |

| | | | | | | |
|---|---|---|---|---|---|---|
| | | | | | 8-14 days since symptom onset PPA 94.7% | |
| | Ellume COVID-19 Home Test | LFA, Fluorescence | Nucleocapsid protein antigen | $10^{3.80}$ $TCID_{50}$/mL | Nasopharyngeal swab samples Within 7 days since symptom onset PPA 96% | 14 |
| | QuickVue SARS Antigen Test | LFA | Nucleocapsid protein antigen | 7.57 x$10^3$ $TCID_{50}$/mL | Nasopharyngeal swab samples Within 5 days since symptom onset PPA 96.6 Negative Percent Agreement (NPA) 99.3 | 15 |
| | Abbott BinaxNOW COVID-19 Ag Card | LFA | Nucleocapsid protein antigen | 140.6 $TCID_{50}$/mL | Nasopharyngeal swab samples 7 days since symptom onset PPA 84.6% 8-10 days since symptom onset PPA 81.9% 11-14 days since symptom onset PPA 78.3% | 16 |
| | Clip COVID Rapid | Lateral flow immunolu | Nucleocapsid | 0.88 x $10^2$ $TCID_{50}$/mL | Nasopharyngeal swab samples | 17 |

| | | | | | | |
|---|---|---|---|---|---|---|
| | Antigen Test | minescent assay | protein antigen | | 5 days since symptom onset<br>PPA 96.9%<br>NPA 100% | |
| | Sampinute COVID-19 Antigen MIA | Magnetic Force-assisted Electrochemical Sandwich Immunoassay (MESIA) | Spike protein antigen | $3.0 \times 10^1$ $TCID_{50}$/mL | Nasopharyngeal swab samples<br>5 days since symptom onset<br>Sensitivity 94.4%<br>Specificity 100% | 18 |
| | Sofia 2 Flu + SARS Antigen FIA | Lateral Flow, Fluorescence, Instrument Read, Multi-Analyte | Nucleocapsid protein antigen | 91.7 $TCID_{50}$/mL | Nasal swab samples<br>5 days since symptom onset<br>PPA 95.2%<br>NPA 100% | 19 |
| | Sofia SARS Antigen FIA | Lateral Flow, Fluorescence | Nucleocapsid protein antigen | $1.13 \times 10^2$ $TCID_{50}$/mL | Nasal swab samples<br>7 days since symptom onset<br>PPA 96.7%<br>NPA 100% | 20 |
| **Emerging SARS-CoV-2 Antigen Tests** | Half-strip LFA | LFA | Nucleocapsid Antigen | 0.65 ng/mL<br>In buffer | Not available | 21 |
| | The SARS-COV-2 Antigen Quantitative Detection Kit (ELISA) (Lot Number 20200508) | ELISA | Serum SARS-COV-2 Nucleocapsid Protein | <1.85 pg/mL | Sensitivity and specificity were 92% (95% confidence interval 81.16–96.85%) and 96.84% (95% confidence interval 95.17–97.15%), respectively | 22 |

| | | | | | | |
|---|---|---|---|---|---|---|
| | Developed by BIOHIT Healthcare (Hefei) Co., Ltd. | | | | | |
| | Bioelectric Recognition Assay (BERA) | Engineer mammalian Vero cells with electroinserting the human chimeric spike S1 antibody. Read the hyperpolarization of the engineered cell membrane with bioelectric/bioelectrochemical sensor. | SARS-CoV-2 S1 spike protein | 1 fg/mL | Need clinical validation | 23 |
| | SARS-CoV-2 RapidPlex | Multiplexed, portable, wireless electrochemical immunoassay on laser-engraved graphene (LEG) electrodes | SARS-CoV-2 nucleocapsid protein (NP), specific immunoglobulins (Igs) against SARS-CoV-2 spike protein (S1) (S1-IgM | Not available | a total of 17 COVID-19 RT-PCR-tested serum samples (10 positive, 7 negative) were assayed, and a total of 8 COVID-19 RT-PCR-tested saliva samples (5 positive, 3 negative) were analyzed | 24 |

| | | | | | | |
|---|---|---|---|---|---|---|
| | | | and S1-IgG), and CRP in blood and saliva | | | |
| | COVID-19 FET sensor | Graphene-based field-effect transistor (FET) biosensing device functionalized with SARS-CoV-2 spike antibody | SARS-CoV-2 spike protein | 1 fg/mL in phosphate-buffered saline and 100 fg/mL clinical transport medium $1.6 \times 10^{1}$ pfu/mL in culture medium, $2.42 \times 10^{2}$ copies/mL in diluted clinical samples | Tested in 3 Nasopharyngeal Swab clinical samples | 25 |
| | eCovSens | A potentiostat based sensor was fabricated using screen printed carbon electrode (SPCE) and immobilized with nCovid-19 monoclonal antibody (nCovid-19Ab) to measure change in | SARS-CoV-2 spike protein | 10 fM in buffer and 90 fM spiked in saliva | Need clinical validation | 26 |

| | | | | | | |
|---|---|---|---|---|---|---|
| | | the electrical conductivity. | | | | |
| | A DNA Aptamer Based Method for Detection of SARS-CoV-2 Nucleocaps id Protein | Aptamer based immunoassay | Serum SARS-COV-2 Nucleocapsid Protein | Not available | Need clinical validation | 27 |
| | Rapid detection method for SARS-CoV-2 S protein using the SARS-CoV-2 receptor ACE2 | Lateral flow immunoassay using the SARS-CoV-2 receptor ACE2 with commercially available antibodies | SARS-CoV-2 spike 1 (S1) protein | 1.86 × $10^5$ copies/mL in nasal swabs | Tested with nasal swabs from COVID-19 patient (n = 4) and healthy subjects (n = 4). Sensitivity 75%, specificity 100%. | 28 |
| | SARSCoV-<br>2 FIC Assay | Fluorescence immunochromatographic (FIC) assay | SARSCoV-<br>2 nucleocapsid protein | Not available | Tested with 251 participants. the sensitivity, specificity and percentage agreement of the FIC assay was 75.6% (95% confidence interval, 69.0e81.3), 100% (95% confidence interval, 91.1e100) and | 29 |

| | | | | | | |
|---|---|---|---|---|---|---|
| | | | | | 80.5% (95% confidence interval, 75.1e84.9) respectively. | |
| | Ultrasensitive SARS-CoV-2 Simoa antigen Assay in Plasma | Single Molecule Array (Simoa) assays | SARS-CoV-2 spike, S1 subunit, and nucleocapsid antigens in the plasma | Detect S1, spike, and N antigens with limits of detection (LOD) of 5 pg/mL (0.07 pmol/L), 70 pg/mL (0.39 pmol/L), and 0.02 pg/mL (0.4 fmol/L), respectively in buffer | SARS-CoV-2 S1 and N antigens were detectable in 41 out of 64 COVID-19 positive patients. In these patients, full antigen clearance in plasma was observed a mean ±95% CI of 5±1 days after seroconversion. | 30 |
| **Emerging SARS-CoV-2 Nucleic Acid Tests** | hybrid capture fluorescence immunoassay (HC-FIA) | LFA | RNA open reading frame 1ab (ORF1ab), envelope protein (E) and the nucleocapsid (N) regions | 500 copies/ml throat swab samples; 1,000 TU/ml for pseudoviruses | Throat swab samples Sputum samples sensitivity 100% specificity 99% | 31 |
| | lateral flow strip membrane | RT-PCR LFA | RNA RdRp gene | 10 copies/test | Swab samples Sputum samples | 32 |

| | | | | | | |
|---|---|---|---|---|---|---|
| | (LFSM) assay | | ORF3a gene<br>N gene | (110 μL)<br>In buffer | Positive agreement 100%<br>Negative agreement 99% | |
| | DETECTR (RT–LAMP/Cas12) assay | CRISPR<br>LAMP<br>LFA | RNA<br>E gene<br>N gene | 500 copies/μL<br>In buffer | Swab samples<br>Positive agreement 95%<br>Negative agreement 100% | 33 |
| | CRISPR/Cas9-mediated triple-line lateral flow assay (TL-LFA) | CRISPR<br>RT-RPA<br>LFA | RNA<br>ORF1ab gene<br>E gene | 25 copies/μL<br>In buffer | Swab samples<br>Negative predictive agreement 100%<br>Positive predictive agreement 97.14% | 34 |
| | iSCAN (in vitro Specific CRISPR-based Assay for Nucleic acids detection) | CRISPR<br>RT-LAMP<br>LFA | RNA<br>E gene<br>N gene | 10 copies/reaction (100 μL)<br>In buffer | Swab samples<br>E gene target:<br>Sensitivity 38%<br>Specificity 100%<br>N gene target:<br>Sensitivity 86%<br>Specificity 100% | 35 |
| **Commercially Available ELISA kits for SARSCoV-2 nucleocapsid protein** | SinoBio SARS-CoV-2 (2019-nCoV) Nucleocapsid Detection ELISA Kit Cat: KIT40588 | ELISA | SARSCoV-2 nucleocapsid protein | 46.68 pg/mL in buffer | Need validation | 36 |
| | Cliniscienc es | ELISA | SARSCoV- | 37.5pg/ml in buffer | Need validation | 37 |

| | | | | | | |
|---|---|---|---|---|---|---|
| | COVID-19 nucleoprotein ELISA Kit Cat# EU3124 | | 2 nucleocapsid protein | | | |
| | Abbexa SARS-CoV-2 Nucleocapsid Protein ELISA Kit Catalogue No: abx365030 | ELISA | SARSCoV-2 nucleocapsid protein | 120 pg/ml in buffer | Need validation | 38 |
| | Ray Biotech COVID-19 N-Protein ELISA CODE: ELV-COVID19N-1 | ELISA | SARSCoV-2 nucleocapsid protein | 70 pg/ml in buffer | Need validation | 39 |
| | ProteinTech SARS-CoV-2 N protein ELISA Kit Cat no : KE30007 | ELISA | SARSCoV-2 nucleocapsid protein | 38 pg/ml in buffer | Need validation | 40 |

### References

1. Chen, H.; Li, Z.; Zhang, L.; Sawaya, P.; Shi, J.; Wang, P., Quantitation of Femtomolar-Level Protein Biomarkers Using a Simple Microbubbling Digital Assay and Bright-Field Smartphone Imaging. *Angewandte Chemie* **2019,** *131* (39), 14060-14066.
2. Everett, J.; Hokama, P.; Roche, A. M.; Reddy, S.; Hwang, Y.; Kessler, L.; Glascock, A.; Li, Y.; Whelan, J. N.; Weiss, S. R.; Sherrill-Mix, S.; McCormick, K.; Whiteside, S. A.; Graham-Wooten, J.; Khatib, L. A.; Fitzgerald, A. S.; Collman, R. G.; Bushman, F., SARS-CoV-2 Genomic Variation in Space and Time in Hospitalized Patients in Philadelphia. *mBio* **2021,** *12* (1).
3. REED, L. J.; MUENCH, H., A SIMPLE METHOD OF ESTIMATING FIFTY PER CENT ENDPOINTS12. *American Journal of Epidemiology* **1938,** *27* (3), 493-497.
4. Hackett, B. A.; Dittmar, M.; Segrist, E.; Pittenger, N.; To, J.; Griesman, T.; Gordesky-Gold, B.; Schultz, D.; Cherry, S., Sirtuin Inhibitors Are Broadly Antiviral against Arboviruses. *mBio* **2019,** *10*.
5. Canny, J., A computational approach to edge detection. *IEEE Trans Pattern Anal Mach Intell* **1986,** *8* (6), 679-98.
6. Duda, R. O. a. H., P. E., Use of the Hough Transformation to Detect Lines and Curves in Pictures. *Comm. ACM* **1972,** *15*, 11-5.
7. A., B. G. a. K., OpenCV. *Dr. Dobb's Journal of Software Tools* **2000,** *3*.
8. US Food and Drug Administration, Coronavirus Disease 2019 (COVID-19) Emergency Use Authorizations for Medical Devices/ In Vitro Diagnostics EUAs. **2020**.

9. CareStart™ COVID-19 Antigen. https://accessbiodiagnostics.net/carestart-covid-19-antigen/
10. LumiraDx SARS-CoV-2 Ag Test. https://www.lumiradx.com/us-en/what-we-do/diagnostics/test-technology/antigen-test
11. BD Veritor System for Rapid Detection of SARS-CoV-2. https://www.bd.com/en-us/offerings/capabilities/microbiology-solutions/point-of-care-testing/bd-veritor-plus-system-for-rapid-covid-19-sars-cov-2-testing
12. VITROS Immunodiagnostic Products SARS-CoV-2 Antigen Reagent Pack. https://www.orthoclinicaldiagnostics.com/global/covid19/antigen-test?utm_source=google&utm_medium=cpc&utm_campaign=G_Ortho_NA_Subbrand_VITROS&utm_content=Ortho_Subbrand_VITROS-BMM&utm_term=+vitros&utm_source=google&utm_medium=cpc&utm_campaign=G_Ortho_Antigen_NA_Subbrand_VITROS&utm_term=%2Bvitros&utm_content=Ortho_Subbrand_VITROS-BMM&gclid=CjwKCAiAsOmABhAwEiwAEBR0ZiT0uB7DDm7knMoUkYC9yg7ifNcYHhhlwzTNbNOVxss0PvP7G2dq8hoC9EsQAvD_BwE
13. Simoa SARS-CoV-2 N Protein Antigen Test. https://www.quanterix.com/simoa-assay-kits/sars-cov-2-n-protein-antigen/
14. Ellume COVID-19 Home Test. https://www.ellumehealth.com/products/consumer-products/covid-home-test
15. QuickVue SARS Antigen Test. https://www.quidel.com/immunoassays/quickvue-sars-antigen-test

16. BinaxNOW COVID-19 Ag Card. https://www.globalpointofcare.abbott/en/support/product-installation-training/navica-brand/navica-binaxnow-ag-training.html
17. Clip COVID Rapid Antigen Test. https://luminostics.com/
18. Sampinute COVID-19 Antigen MIA. https://www.celltrion.com/en-us/kit/sampinute
19. Sofia 2 Flu + SARS Antigen FIA. https://www.quidel.com/immunoassays/sofia-2-flu-sars-antigen-fia
20. Sofia SARS Antigen FIA. https://www.quidel.com/immunoassays/rapid-sars-tests/sofia-sars-antigen-fia
21. Grant, B. D., Anderson, C. E., Williford, J. R., et al. (2020). SARS-CoV-2 coronavirus nucleocapsid antigen-detecting half-strip lateral flow assay toward the development of point of care tests using commercially available reagents. *Analytical chemistry*, *92*(16), 11305-11309.
22. Li, T.; Wang, L.; Wang, H.; Li, X.; Zhang, S.; Xu, Y.; Wei, W., Serum SARS-COV-2 Nucleocapsid Protein: A Sensitivity and Specificity Early Diagnostic Marker for SARS-COV-2 Infection. *Frontiers in Cellular and Infection Microbiology* **2020,** *10* (470).
23. Mavrikou, S.; Moschopoulou, G.; Tsekouras, V.; Kintzios, S., Development of a Portable, Ultra-Rapid and Ultra-Sensitive Cell-Based Biosensor for the Direct Detection of the SARS-CoV-2 S1 Spike Protein Antigen. *Sensors (Basel)* **2020,** *20* (11), 3121.
24. Torrente-Rodríguez, R. M.; Lukas, H.; Tu, J.; Min, J.; Yang, Y.; Xu, C.; Rossiter, H. B.; Gao, W., SARS-CoV-2 RapidPlex: A Graphene-Based Multiplexed Telemedicine Platform for Rapid and Low-Cost COVID-19 Diagnosis and Monitoring. *Matter* **2020,** *3* (6), 1981-1998.
25. Seo, G.; Lee, G.; Kim, M. J.; Baek, S.-H.; Choi, M.; Ku, K. B.; Lee, C.-S.; Jun, S.; Park, D.; Kim, H. G.; Kim, S.-J.; Lee, J.-O.; Kim, B. T.; Park, E. C.; Kim, S. I., Rapid Detection of COVID-19 Causative Virus (SARS-CoV-2) in Human Nasopharyngeal Swab Specimens Using Field-Effect Transistor-Based Biosensor. *ACS Nano* **2020,** *14* (4), 5135-5142.
26. Mahari, S.; Roberts, A.; Shahdeo, D.; Gandhi, S., eCovSens-Ultrasensitive Novel In-House Built Printed Circuit Board Based Electrochemical Device for Rapid Detection of nCovid-19 antigen, a spike protein domain 1 of SARS-CoV-2. *bioRxiv* **2020**, 2020.04.24.059204.
27. Chen, Z.; Wu, Q.; Chen, J.; Ni, X.; Dai, J., A DNA Aptamer Based Method for Detection of SARS-CoV-2 Nucleocapsid Protein. *Virologica Sinica* **2020,** *35* (3), 351-354.
28. Lee, J.-H.; Choi, M.; Jung, Y.; Lee, S. K.; Lee, C.-S.; Kim, J.; Kim, J.; Kim, N. H.; Kim, B.-T.; Kim, H. G., A novel rapid detection for SARS-CoV-2 spike 1 antigens using human angiotensin converting enzyme 2 (ACE2). *Biosensors and Bioelectronics* **2021,** *171*, 112715.
29. Diao, B.; Wen, K.; Zhang, J.; Chen, J.; Han, C.; Chen, Y.; Wang, S.; Deng, G.; Zhou, H.; Wu, Y., Accuracy of a nucleocapsid protein antigen rapid test in the diagnosis of SARS-CoV-2 infection. *Clinical microbiology and infection : the official publication of the European Society of Clinical Microbiology and Infectious Diseases* **2020**.
30. Ogata, A. F.; Maley, A. M.; Wu, C.; Gilboa, T.; Norman, M.; Lazarovits, R.; Mao, C.-P.; Newton, G.; Chang, M.; Nguyen, K.; Kamkaew, M.; Zhu, Q.; Gibson, T. E.; Ryan, E. T.; Charles, R. C.; Marasco, W. A.; Walt, D. R., Ultra-Sensitive Serial Profiling of

SARS-CoV-2 Antigens and Antibodies in Plasma to Understand Disease Progression in COVID-19 Patients with Severe Disease. *Clinical Chemistry* **2020,** *66* (12), 1562-1572.

31. Wang, D., He, S., Wang, X., et al. (2020). Rapid lateral flow immunoassay for the fluorescence detection of SARS-CoV-2 RNA. *Nature biomedical engineering*, 1-9.

32. Yu, S., Nimse, S. B., Kim, J., Song, K. S., & Kim, T. (2020). Development of a Lateral Flow Strip Membrane Assay for Rapid and Sensitive Detection of the SARS-CoV-2. *Analytical chemistry*, *92*(20), 14139-14144.
33. Broughton, J. P., Deng, X., Yu, G., et al. (2020). CRISPR–Cas12-based detection of SARS-CoV-2. *Nature Biotechnology*, 1-5.
34. Xiong, E., Jiang, L., Tian, T., et al. (2020). Simultaneous dual-gene diagnosis of SARS-CoV-2 based on CRISPR/Cas9-mediated lateral flow assay. *Angewandte Chemie*.
35. Ali, Z., Aman, R., Mahas, A., Rao, G. S., et al. (2020). iSCAN: An RT-LAMP-coupled CRISPR-Cas12 module for rapid, sensitive detection of SARS-CoV-2. *Virus research*, *288*, 198129.
36. https://www.sinobiological.com/elisa-kits/cov-nucleocapsid-kit40588

37. https://www.clinisciences.com/en/sars-cov-2-elisa-kits-nucleocapsid-5143/covid-19-nucleoprotein-elisa-kit-565019800.html

38. https://www.abbexa.com/sars-cov-2-nucleocapsid-protein-elisa-kit

39. https://www.raybiotech.com/covid-19-n-protein-elisa-en/
40. https://www.ptglab.com/products/Human-SARS-CoV-2-N-protein-ELISA-Kit-KE30007.htm
